# Evaluation of seven different rapid methods for nucleic acid detection of SARS-COV-2 virus

**DOI:** 10.1101/2021.04.15.21255533

**Authors:** Sally A. Mahmoud, Esra Ibrahim, Subhashini Ganesan, Bhagyashree Thakre, Juliet George Teddy, Preeti Raheja, Walid Abbas Zaher

## Abstract

**Background:** In the current COVID-19 pandemic there is mass screening of SARS-CoV-2 happening round the world due to the extensive spread of the infections. There is a high demand for rapid diagnostic tests to expedite identification of cases and to facilitate early isolation and control spread. Hence this study evaluates seven different rapid nucleic acid detection assays that are commercially available for SARS-CoV-2 virus detection.

**Methods:** Nasopharyngeal samples were collected from 4859 participants and were tested for SARS-CoV-2 virus by the gold standard RT-PCR method along with one of these seven rapid methods of detection. Evaluation of the rapid nucleic acid detection assays was done by comparing the results of these rapid methods with the gold standard RT-qPCR results for SARS-COV-2 detection.

**Results:** AQ-TOP had the highest sensitivity (98%) and strong kappa value of 0.943 followed by Genechecker and Abbot ID NOW. The POCKIT (ii RT-PCR) assay had the highest test accuracy of 99.29% followed by Genechecker and Cobas Liat. Atila iAMP showed the highest percentage of invalid reports (35.5%) followed by AQ-TOP with 6% and POCKIT with 3.7% of invalid reports.

**Conclusion:** Genechecker system, Abbott ID NOW and Cobas Liat, were found to have best performance and agreement when compared to the standard RT-PCR for COVID-19 detection. With further research, these rapid tests have the potential to be employed in large scale screening of COVID-19.

## Background

The COVID-19 pandemic has affected more than 112 million people worldwide and continues to spread and remains a public health challenge. [1] The real-time reverse-transcription PCR (RT-PCR) remains the gold standard for testing SARS-COV-2 and is approved by both the WHO and CDC. But this method requires a well-established lab set up, expensive instruments, well trained and skilled manpower and long hours. In the current scenario of the pandemic, with such large numbers being affected conducting these tests with limited lab capacities is challenging. [2] The lab facilities are overburdened, and molecular testing is time consuming which delays reporting and that in turn limits the containment of spread. [3] Hence there is a demand for alternative testing strategies that are rapid and less sophisticated. [4].

In SARS-CoV-2 human-to-human transmission is through droplets or direct contact [5] and its symptoms are very similar to flu, hence molecular tests are critical to differentiate and detect COVID-19 infections. In the early stages of infection, the viral load is usually high in patients and studies show that a single swab can contain more than a million viral particles,[6] hence nucleic acid testing is the most efficient form of testing in the early stages, and identifying infections earlier is vital. As mass testing, early detection and isolation are crucial for containing the spread of infection, evaluation of these rapid tests becomes imperative.

Currently there are many nucleic acid detection assays that have obtained emergency authorization by FDA to detect SARS-COV-2 and have been largely implemented around the globe. The study compares seven different molecular tests that detects nuclear RNA of SARS-COV-2 and evaluates them against the standard RT-PCR.

## Materials and Methods

This study obtained ethical approval from the IRB of Department of Health (DOH), Abu Dhabi. The study was conducted among people who were admitted in the COVID-19 field hospital and among people who were getting screened for COVID-19 infections in the laboratory. After getting informed consent from the participants nasopharyngeal swabs (NPS) were collected from the study participants. For the wet swab, a sterile swab was used for collection and then was placed into the universal transport medium (UTM). An additional dry swab was collected from participants who were willing to give an additional sample. All samples that were collected outside the lab were transported to the lab immediately and samples that were not processed immediately were stored at -80 degree Celsius.

A total of 5181 participants were tested for SARS-CoV-2 virus by the gold standard RT-PCR method for SARS-CoV-2 detection and with one of these seven rapid methods of detection. The rapid tests were evaluated by comparison the rapid nucleic acid detection assays results with the standard RT-PCR results for SARS-COV-2 detection. Of these seven rapid detection methods, two were based on loop isothermal nucleic acid amplification technology (Abbott ID NOW, AQ-TOP), one on OMEGA amplification (Atila iAMP), one direct RT-PCR without RNA extraction method (Direct NPS), one based on insulated isothermal polymerase chain reaction (iiPCR) technology, one on automated multiplex real-time RT-PCR assay (Cobas Liat) and one based on microfluidic chip-based PCR method (Genechecker).

### The seven rapid nucleic acid detection assays

#### The Abbott ID NOW COVID-19 assay [7]

689 samples were tested using this method. It is an automated assay that utilizes isothermal nucleic acid amplification technology for the qualitative detection of nucleic acid from the SARS-CoV-2 virus from nasopharyngeal swabs from individuals who are suspected of COVID-19. It detects the RNA-dependent RNA polymerase (RdRp) gene segment of SARS-CoV-2 and is performed on the ID NOW instrument. Fluorescently labeled molecular beacons are used to specifically identify each of the amplified RNA targets. The testing was performed as per manufacturer’s instructions. It takes 1-2 minutes active preparation and the time to result is about 13 minutes. It has received FDA emergency authorization for use. [8] In this study, the testing was done onsite for detecting SARS-COV-2 from a dry swab collected from the participants. Results are directly displayed on the screen as positive or negative along with the internal control validity.

#### Atila iAMP COVID-19 detection [9]

197 samples were tested using this method, which is a real-time reverse transcription based on a proprietary isothermal amplification technology termed OMEGA amplification. OMEGA primer sets are designed to specifically detect RNA and later cDNA from the N and ORF-1ab genes of the SARSCoV-2 virus in nasal, nasopharyngeal and/or oropharyngeal swabs from individuals who are suspected of COVID-19. The iAMP COVID-19 assay has the ability to detect SARS-CoV-2 RNA directly from samples without prior RNA extraction process. All procedures as per manufacturer instructions were followed. The turnaround time is 60 mins. It has received FDA emergency authorization for use.[10] In this method, the control used is a human gene that is amplified and measured in the HEX channel. If a sample shows no exponential amplification curve in the HEX channel but an exponential curve in the FAM channel, the sample is still reported as a valid run and will be interpreted according to maufacturers instructions. If there is no exponential amplification curve in any channel, the sample test result is termed invalid.

#### AQ-Top Plus COVID-19 Rapid Detection Kit

[11] 226 samples were run through this kit. It is a Real-Time Loop Mediated Isothermal Amplification (RT-LAMP) test intended for the qualitative detection of nucleic acid from SARS-CoV-2 in clinical respiratory specimens like nasopharyngeal swabs. The AQ-TOP COVID-19 Rapid Detection Kit PLUS uses dual-labeled Peptide Nucleic Acid (PNA) probes that target ORF1ab and N gene for detection of SARS-CoV-2 RNA. It has a fast turnaround time of 20 minutes. It has also received FDA emergency authorization for use.[12] The results are interpreted as per manufacturer’s instructions. If no amplification curves are detected in both the FAM and VIC fluorescent channels or if an amplification curve is detected but at a CT more than 30 the test is considered invalid.

#### Direct Nasopharyngeal Swab (Direct NPS) Realtime RT-PCR [13]

200 samples were tested by this method. The NPS samples collected were stored in the lab at -80 degree Celsius and was run after few days. It is an RNA-extraction free, RT-qPCR method for SARS-CoV-2 detection.

The direct NPS method was tested with various transport media for direct lysis. Bio-Speedy Direct RT-qPCR SARS-Cov-2 kit with the vNAT buffer contains guanidinium thiocyanate and was validated with 50 samples. 90 samples were used to validate DIAGNOVITAL Magicprep fast RNA isolation kit and 10 samples were validated with OriginSafe Viral Lysis Transport Kit, which contains Viral Lysis Transport Medium (OriginSafe VLTM) that disrupts lipid membrane and inactivates nucleases. All 3 kits were unsuccessful to produce a good quality extract and all tests presented invalid results. However, direct lysis using proteinase K was relatively more successful and was pursued for evaluation. Nasal Swabs are treated with proteinase K followed by a heat inactivation step and is then directly added to the master mix for RT-qPCR. For best results, optimization of the ratio of proteinase K and the sample mixture was done in the lab, testing five different ratios starting from 5ul of the proteinase K with 50 ul of the sample sequentially to 40ul of proteinase K and 15ul of sample mixture and the best correlation with the reference values were attained at 15 ul of proteinase K with 40 ul sample. With this optimization further testing was done, turnaround time is around 2 hours.

#### Genechecker PCR system-UF 300 – RT PCR system [14]

1133 samples were tested in this method. It is an innovative microfluidic chip-based PCR method with a compact device and ultra-fast ramping of 40 cycles in 20 minutes through a unique thermal cycling mechanism. The kit has been developed to detect both RdRP gene and N gene of SARS-COV-2. Two fluorescent channels are used, FAM and ROX to detect the gene and to serve as an internal control respectively. The primer pairs and probes for the detection of these two target genes are pre-labeled in the wells of the test chip so that user does not have to pipette primers and probes for running the test. Four samples can be tested per run and the standard turnaround time is 45 minutes with 10 – 15 minutes of preparation.

The results are displayed on screen as either N gene and RdRP gene detected or not detected along with the control validity. This makes the reporting easier, and it also displays the amplification curves for easier reading. The positive control should be positive, and template control should be negative to confirm the test validity, if not the test is reported invalid.

#### Cobas Liat system SARS-CoV-2 & Influenza A/B nucleic acid test [15]

524 samples were tested with this method. It is an automated multiplex real-time RT-PCR assay for in-vitro qualitative detection and differentiation of SARS-CoV-2, influenza A, and influenza B virus RNA in nasopharyngeal and nasal swabs.The assay targets both the ORF1 a/b non-structural region and nucleocapsid protein gene that are unique to SARS-CoV-2. An Internal Process Control (IPC) is present to control for adequate processing of the target virus through steps of sample purification, nucleic acid amplification, and to monitor the presence of inhibitors in the RT-PCR processes. The results are displayed as SARS-CoV-2 detected, not detected or invalid within 20 minutes. It has got FDA emergency authorization for use. [16]

#### POCKIT SARS-CoV-2 (orf lab) (RT-ii PCR) assay [17]

It is a PCR test for qualitative detection of SARS-CoV-2 from nasopharyngeal swabs and 2212 samples were tested with this method. It uses the insulated isothermal polymerase chain reaction (iiPCR) technology and must be used with a POCKIT Central Nucleic Acid Analyzer. It generates fluorescent signals, 520 nm and 550 nm, when the specific nucleic acid sequences of SARS-CoV-2 and the internal Control are amplified, respectively. When both the 520 nm and 550 nm signals are positive the sample is interpreted positive for SARS-COV-2 and when 520 nm is negative, and 550 nm is positive the sample is termed negative for SARS-COV-2 virus. When both signals are negative or not detected it is termed invalid.

### Statistical analysis

The results of the standard RT – qPCR was used as a standard reference and the results obtained from the rapid methods were compared to the standard reference reports.

Sensitivity, specificity, positive predictive value (PPV), negative predictive value (NPV) and accuracy were calculated for each method. Cohen’s kappa values were calculated to measure agreement, along with percent positive agreement (PPA), negative agreement percent (NPA) and overall agreement were calculated. SPSS statistical software was used for all statistical analysis.

## Results

The sensitivity, specificity, positive predictive value (PPV), negative predictive value (NPV) and accuracy were calculated for the seven rapid methods by comparing with the standard RT-PCR results. The results are shown in figure 1. (Figure 1)

**Figure 1:**
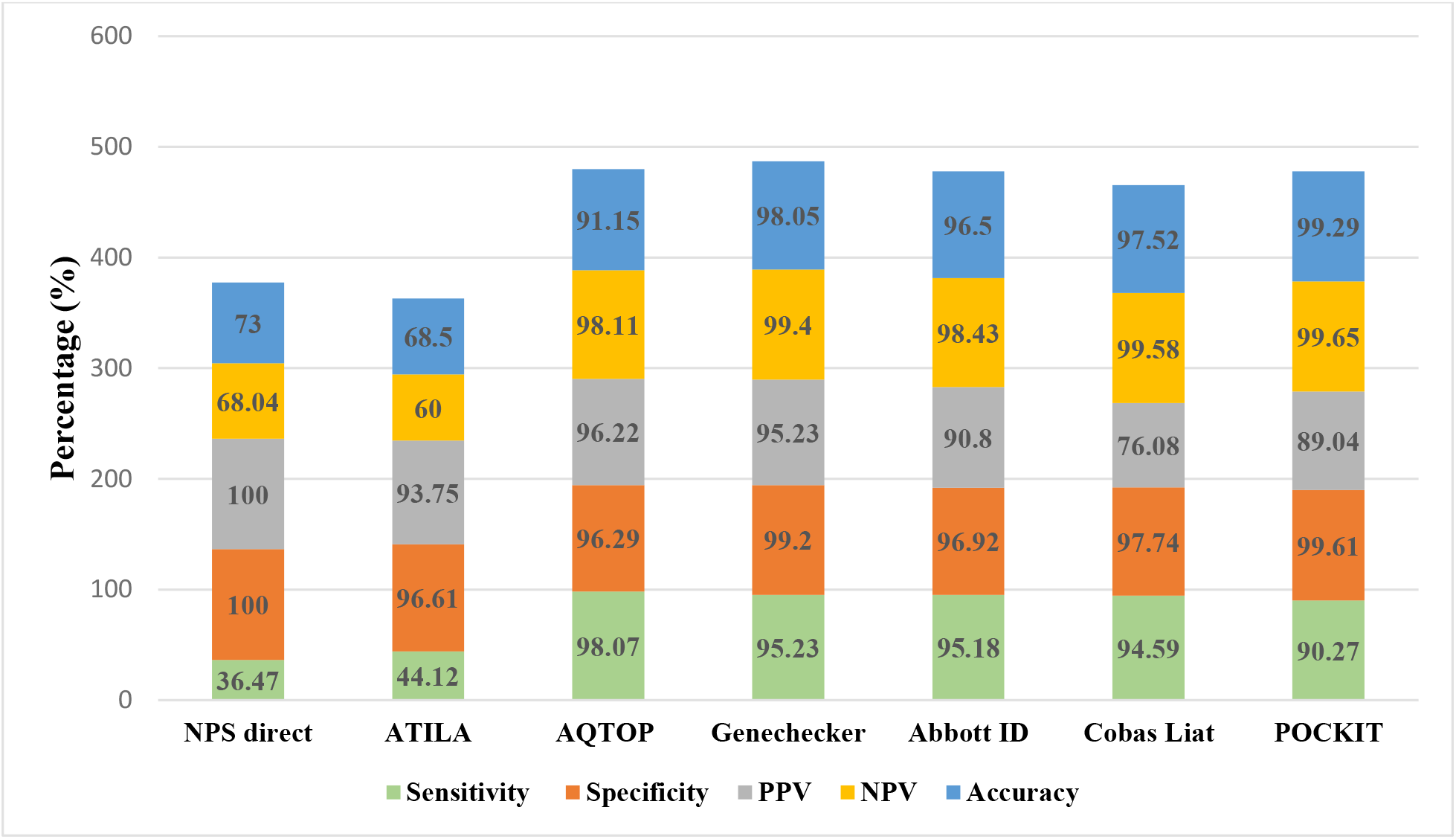
Comparison of the seven rapid methods of nucleic acid detection.

AQ-TOP and Genechecker had the highest sensitivity and positive predictive value followed by sensitivity of Abbott ID NOW and Cobas Liat. The Direct NPS RT-qPCR method and Atila had the lowest sensitivity. The POCKIT had the highest accuracy followed by Genechecker and Cobas Liat.

The positive, negative, and overall agreement percentage along with Cohens’ kappa value were calculated for the seven methods and are presented in Table 1.

**Table 1:** Shows the agreement of the rapid tests results with the standard reference test results.

| Molecular assays (n) | Standard reference test |  | Kappa (k) (p value) | Positive agreement (%) | Negative agreement (%) | Overall Agreement (%) |
| --- | --- | --- | --- | --- | --- | --- |
|  | Positive | Negative |  |  |  |  |
| Direct NPS RT-PCR (200) |  |  |  |  |  |  |
| Positive | 31 | 0 | 0.398 | 36.47 | 100 | 73 |
| Negative | 54 | 115 | (<0.001) |  |  |  |
| Atila iAMP (127) |  |  |  |  |  |  |
| Positive | 30 | 2 | 0.391 | 44.11 | 96.61 | 68.50 |
| Negative | 38 | 57 | (<0.001) |  |  |  |
| AQ – Top (212) |  |  |  |  |  |  |
| Positive | 102 | 4 | 0.943 | 98.07 | 96.29 | 91.15 |
| Negative | 2 | 104 | (<0.001) |  |  |  |
| **Genechecker**<br>**(1128)** |  |  |  |  |  |  |
| Positive | 120 | 8 | 0.938 | 95.23 | 99.20 | 98.75 |
| Negative | 6 | 994 | (<0.001) |  |  |  |
| **Abbott ID**<br>**NOW (686)** |  |  |  |  |  |  |
| Positive | 158 | 16 | 0.906 | 95.18 | 96.92 | 96.50 |
| Negative | 8 | 504 | (<0.001) |  |  |  |
| **Cobas Liat**<br>**(524)** |  |  |  |  |  |  |
| Positive | 35 | 11 | 0.830 | 94.59 | 97.74 | 97.52 |
| Negative | 2 | 476 | (<0.001) |  |  |  |
| **POCKIT**<br>**(2131)** |  |  |  |  |  |  |
| Positive | 65 | 8 | 0.893 | 90.27 | 99.61 | 99.29 |
| Negative | 7 | 2051 | (<0.001) |  |  |  |

The positive percentage agreement with the standard RT-qPCR was strongest with AQ-Top followed by Genechecker. Similarly, the negative agreement was best with POCKIT, Cobas Liat and Genechecker. AQTOP. The overall agreement was highest in POCKIT and the overall agreement percentage was poor in Direct PCR and Atila iAMP.

The number of positive cases missed, and details of the results reported invalid by the rapid tests are shown in Table 2. Atila iAMP showed the highest percentage of invalid reports missing a large proportion (44.7%) of positive cases followed by AQ-TOP with 7% invalid reports. The Direct NPS-RT-PCR and Cobas Liat did not show any invalid reports.

**Table 2:** Invalid reports and positive cases missed by the rapid methods.

| **Rapid methods (n)** | **Invalid reports**<br>**(n(%))** | **Positive cases reported**<br>**by standard RT-qPCR**<br>**missed by the rapid**<br>**tests (n (%))** |
| --- | --- | --- |
| Direct Nasopharyngeal Swab Realtime RT-PCR (200) | 0 | 0 |
| Atila iAMP COVID- 19 detection (197) | 70 (35.5) | 55 (44.7) |
| AQ-TOP COVID-19 Rapid Detection (226) | 14 (6.1) | 3 (2.8) |
| Genechecker- UF 300 – RT PCR system (1133) | 5 (0.4) | 5 (3.1) |
| The Abbott ID NOW COVID-19 assay (689) | 3 (0.4) | 2 (1.2) |
| Cobas Liat (524) | 0 | 0 |
| POCKIT SARS-CoV-2 (orf lab) Premix<br>Reagent (2212) | 81 (3.7) | 10(1.3) |

## Discussion

In this ongoing COVID-19 pandemic with extensive transmission of infection, rapid molecular tests are very crucial for early identification and isolation. The molecular assays evaluated in this study are all rapid with low complexity thus requires less hands-on time, which is the need of the hour hence assays were evaluated in comparison to the standard RT-PCR.

Our study found that the sensitivity of the Atila iAMP assay and Direct lysis RT-PCR method for nasopharyngeal swab RT-qPCR was very low. While some studies have demonstrated no substantial difference in sensitivity for direct NPS method when compared to standard RT-qPCR for molecular detection of SARS-CoV-2 [18], there are also evidences that show, amplification directly from samples can reduce efficiency of PCR. [19]. This can be attributed to the presence of inhibitors that interferes with the polymerase activity and decrease the sensitivity and accuracy of detection. [20] Another reason may be due to inadequate extraction quality in the direct method, further the samples that were used in this study for the direct NPS RT-qPCR method were stored in the lab at -80 degrees Celsius for 3 – 4 days. This might be one of the reasons for low sensitivity as a study done in Germany has shown that direct NPS performs better when the samples are fresh and RNA extraction would be required if the samples are stored longer. [21]

The Atila iAMP assay has been claimed to have around 87% PPA and 100% NPA with the reference RT-PCR assay. [22], but in this study we found that the sensitivity was very low, and it reported a large number of invalid results, missing more than 40 % of positive cases when compared to the standard RT-qPCR results. The agreement percentage (kappa agreement) with the NPS results was low. In this assay, 93 positive samples were reported as either negative or invalid and the mean Ct value of these positive samples were 31.54± 4.84. The test missed weak/ low viral load samples, that had Ct values more than 30. This was supported by the study in Stanford, that showed Atila had lower sensitivity and required high volume of nucleic acid eluate. [22] Further studies have shown that molecular tests using the LAMP technology have reported false negative results in low viral load. [23] However the assay showed high specificity which can be explained by the four different primers that are used in this assay to detect six different sequences of the RNA of SARS-COV-2. [24] Also the test does not require RNA extraction prior to amplification, thus reducing the time to run along with the ability to process 96 samples per run and providing the results within 60 minutes. While there are positive aspects, the sensitivity and reliability of the method are important parameters for screening, hence with lower sensitivity higher percentages of false negative results would be expected and the high percentage of invalid results makes the test less reliable as a rapid method, for these reasons the test cannot be widely employed as a screening test.

In our study the Abbott and AQ-TOP both used the LAMP technology and had high sensitivity, specificity, PPA, NPA and the kappa coefficient was strong. Similar studies have reported LAMP assays to be identical with the RT-PCR tests and reported similar sensitivity and specificity [25] The drawback of TOP - AQ was that it showed 6% invalid reports, which is a relatively high number compared to other tests. But this method is quick with faster amplification and high throughput, 96 samples can be tested in each run, results are easily readable and requires less specialized equipment. It further has the ability for amplification of multiple targets in a single reaction.[26]

The Abbott ID NOW had an accuracy of 96.5%, with sensitivity and specificity comparable to the standard RT – qPCR and lowest percentage of invalid reports compared to other methods. It also showed a strong agreement with the RT-PCR results. A study by Rhoads in the USA reported similar findings, which showed that the assay had a good PPA of 94%. [27] Another study done in New York had shown that when compared to Xpert Xpress, ID NOW revealed good PPA only when sample had low Ct values and the PPA was low in the higher Ct values samples or when the viral load was low. [28] Similar reports were published by another study that validated RT-LAMP, which showed that samples having Ct values less than 30 showed 100% sensitivity, while samples with Ct values more than 35 showed only 54% concordance with the RT-qPCR results.[29] However in our study, we found that the concordance with the RT-qPCR results did not change with the Ct values. Further, the Abbott ID is compact, can be used as a bedside test and it is rapid as it detects the positive results in just 7 minutes and negative results in 13 minutes, although only one sample can be tested per run.

The Genechecker had a high sensitivity and concordance with the RT-qPCR reports, with the least percentage of invalid reports. It has statistically significant 0.938 Cohen’s kappa value and thereby strong agreement with the results of the standard reference test. Further this method has a turnaround time of 45 minutes and 4 samples can be tested per run. But the preparation of the samples requires 5 to 10 minutes and requires manual pipetting which is recommended to be done in a biosafety cabinet. Therefore, though Genechecker is a compact instrument cannot be used as a bedside test. Also for efficient usage four samples need to be run together as there would be wastage of the kit cartridges. There aren’t many studies on the evaluation of Genechecker but our study shows the method has great potential as a rapid detection method.

The Cobas Liat automated RT-PCR showed the PPA of 94.5 % and accuracy of 97.5% with the lowest PPV of 76% and the results can be obtained in 20 minutes . Another study that evaluated the clinical performance of Cobas Liat showed that its accuracy was 98.6%, positive percent agreement was 100% and the negative percent agreement was 97.4%. which was very similar to our study report. [30] However a recent report warns of potential false positives with Cobas rapid test that was alleged to sporadic assay tube leakage or abnormal PCR cycling in the reaction tubes. [31] This might explain the high false positives and lowest PPV observed in our study with Cobas Liat.

The POCKIT SARS-CoV-2 (orf lab) (RT-ii PCR) assay had the highest overall agreement with the reference RT-PCR method. The turnaround time is 85 minutes with a throughput of 8 samples per run. The positive A study that validated its clinical performance comparing it to standard RT-PCR assay also showed that positive agreement was 96.8% and kappa value of 0.93. [32] However in our study the positive agreement was only 90% this might be because the number of positive samples tested were only 75 samples compared to 2137 negative reports. The test reported high number of invalid reports which was 3.7 % of the total samples.

## Strengths and limitations

The study has evaluated commercial assays available in the market for rapid detection of SARS-COV-2, which is the need of the hour. All the results are compared to the gold standard RT-PCR recommended for SARS-COV-2 detection, which gives an insight of the performance of these rapid methods based on the standard comparison.

The limitation is that the number of samples tested with each rapid method varies, which was based on the availability of kits and reagents.

## Conclusion

In summary, the evaluation of seven rapid methods of detection of SARS-COV-2 using NPS shows that the direct (extraction free) NPS method and Atila had low sensitivity, along with the high number of invalid reports with Atila, which made the test unreliable. While AQ-TOP had good sensitivity and rapid turnaround time of 20 minutes but again the percentage of invalid reports were high using this method. The POCKIT had highest overall agreement percentage but with lower sensitivity than most other rapid tests and high number of invalid reports. AQ-TOP, Abbott and GeneChecker had high sensitivity a strong kappa value and rapid turnaround time of 20, 15 and 45 minutes, respectively. Cobas Liat, Abbott ID NOW and Genechecker system had the least number of invalid reports however the throughput for each is 1,1 and 4 respectively. Further studies reviewing of all these parameters are needed, to consider implementing these rapid tests in large scale in detection of SARS-COV-2.

## Data Availability

The data is available with the corresponding author, Dr. Sally, Director of Biogenix G42 lab and will be produced on request.

## Authors contribution

1. Sally A. Mahmoud: conceptualization, Methodology, validation, reviewing and editing, supervision and project administration
2. Esra Ibrahim : Conceptualization, Methodology, validation, reviewing and editing
3. Subhashini Ganesan: Conceptualization, Methodology, writing original draft, validation, reviewing and editing
4. Bhagyashree Thakre: Methodology, validation, reviewing and editing
5. Juliet G Teddy: Methodology, validation, reviewing and editing
6. Preeti Raheja: Methodology, validation, reviewing and editing
7. Walid Z Abbas: Reviewing, editing, supervision and project administration

## Ethics approval and consent to participate

The Ethics approval was obtained from Department of Health (DOH) Institutional review board (IRB), Abu Dhabi. All methods were carried out in accordance with relevant guidelines and regulations. The Department of Health (DOH) Institutional review board (IRB), Abu Dhabi.

## Informed consent statement

Informed consent was obtained from all participants in the study.

